# High Probability of Lynch Syndrome among colorectal cancer patients in Indonesia is associated with higher occurrence of KRAS and PIK3CA mutations

**DOI:** 10.1101/2024.03.03.24303469

**Authors:** Didik Setyo Heriyanto, Naomi Yoshuantari, Gilang Akbariani, Vincent Lau, Hanifa Hanini, Zulfa Hidayati, Muhammad Zulfikar Arief, Andrew Nobiantoro Gunawan, Asep Muhamad Ridwanuloh, Wien Kusharyoto, Adeodatus Yuda Handaya, Mohammad Ilyas, Johan Kurnianda, Susanna Hilda Hutajulu, Susanti Susanti

## Abstract

**Background:** In Indonesia, early-onset colorectal cancer (EOCRC) rates are higher in patients <50 years old compared to western populations, possibly due to a higher frequency of Lynch Syndrome (LS) in CRC patients. We aim to examine the association of KRAS and PIK3CA mutation with LS.

**Methods:** In this cross-sectional study, the PCR-HRM-based test was used for screening of MSI mononucleotide markers (BAT25, BAT26, BCAT25, MYB, EWSR1), MLH1 promoter methylation, and oncogene mutations of BRAF(V600E), KRAS (exon 2 and 3), and PIK3CA (exon 9 and 20) in FFPE DNA samples.

**Results:** All the samples (n=244) were from Dr. Sardjito General Hospital Yogyakarta, Indonesia. KRAS and PIK3CA mutations were found in 151/244 (61.88%) and 107/244 (43.85%) of samples respectively. KRAS and PIK3CA mutations were significantly associated with MSI status in 32/42 (76.19%) and 25/42 (59.52%) of samples respectively. KRAS mutation was significantly associated with LS status in 26/32 (81.25%) of samples. The PIK3CA mutation was present in a higher proportion in LS samples of 19/32 (59.38%), but not statistically significant. Clinicopathology showed that KRAS mutation was significantly associated with right-sided CRC and higher histology grade in 39/151 (25.83%) and 24/151 (16.44%) samples respectively. PIK3CA mutation was significantly associated with female sex and lower levels of TILs in 62/107 (57.94%) and 26/107 (30.23%) samples respectively. KRAS and PIK3CA mutations did not significantly affect overall survival (120 months) in LS and non-LS patients.

**Conclusions:** High probability of LS in Indonesian CRC patients is associated with KRAS and PIK3CA mutations.

## INTRODUCTION

Colorectal cancer (CRC) is the third most prevalent cancer worldwide and as well as one of the most deadly. Approximately in Indonesia, over 35,000 patients are diagnosed with CRC each year [1]. Three provinces in Indonesia have the highest incidence of CRC: Jakarta, Central Java, Yogyakarta. Early-onset colorectal cancer (EOCRC) accounts for nearly 30% of total CRC patients, three times higher than in Europe, the UK, and the USA [2]. The epidemiological data in Indonesia showed that the proportion of CRC patients <40 years old was more than 30% [3]. This incidence in Indonesia was higher in males (54%) than in females (46%), with a peak age of 50–54 years [2].

Lynch syndrome (LS) also known as hereditary non-polyposis colon cancer (HPNCC) is a hereditary type of CRC. This syndrome is characterized by early-onset (<50 years) [4, 5]. Our previous study introduced the higher frequency of LS cases in Yogyakarta, Indonesia which linked to a high risk of EOCRC [6].

Three molecular pathways have been identified for the pathogenesis of CRC: chromosomal instability (CIN), microsatellite instability (MSI), and the CpG Island Methylator Phenotype (CIMP) [7]. The CIN involves gene mutations in APC, KRAS, SMAD4, and TP53, while MSI is caused by mutations in Mismatch Repair (MMR) genes such as MSH2, MLH1, MSH3, PMS1, and PMS2 genes [8]. The CIMP, which is characterized by CG dinucleotide methylation in the promoters of numerous genes, is associated with distinct clinical and pathological attributes in tumors. These subtypes, as described by Jass Classification, include CIMP high/MSI high (12% of CRC), CIMP low/MSI low or microsatellite stable (20%), CIMP negative/microsatellite stable (57%), and Hereditary Non Polyposis Colorectal Cancer (HNPCC), with CIMP negative/MSI high and negative for BRAF mutations [7, 9–12]. Testing for MSI and MMR protein deficiency is commonly the first step in LS diagnostics due to germline mutations of MMR genes. On the contrary, epigenetic silencing of the MLH1 and somatic mutation of BRAF are common in sporadic tumors with MSI but very rarely occur in tumors arising in LS [13, 14].

KRAS mutations, the most common RAS family mutation, affect cell proliferation, differentiation, senescence, and apoptosis in 40% of sporadic CRC. These mutations increase CRC tumor aggressiveness, reduce survival rates, and promote treatment resistance [15]. Anti-EGFR monoclonal antibodies, such as cetuximab and panitumumab, are ineffective in CRC patients with KRAS codon 12, 13, or 14 mutations. Hence, these agents are only effective in RAS wild-type tumors [16]. BRAF mediates RAS-RAF-MAP kinase growth signal responses. BRAF mutations are found in 4% of MSI-low tumors and 40% of MSI-high tumors [17]. The most frequent of these mutations are BRAF^V600E^ (Val600Glu). These BRAF^V600E^ mutations help to differentiate between familial and sporadic CRC and are associated with poorer prognosis. Generally, BRAF mutations are confined to tumors without KRAS exon 2 mutations. BRAF is downstream of activated KRAS in the EGFR pathway, making cetuximab or panitumumab ineffective for inhibiting EGFR, unless given BRAF Inhibitor [10, 18].

Phosphatidylinositol-3-kinase (PI3K) is a heterodimeric lipid kinase involved in cell signaling and cell membrane function. Mutations in PIK3CA have been reported in 10-20% of CRC cases [19]. PIK3CA mutations are associated with worse clinical outcomes and a negative predictor of response to anti-EGFR targeted therapy [20]. It has been shown that RAS mutations are negative predictors of anti-EGFR mAb response and survival benefit [20]. Over 80% of PIK3CA mutations are found in two hotspots: the helicase domain of exon 9 and the kinase domain in exon 20. Several studies have analyzed PIK3CA mutation in these hotspots for the discrepancy of the predictive values of PIK3CA as a biomarker for anti-EGFR [20, 21]. The PI3K pathway’s downstream effectors include AKT and mTOR, which increase cell cycle regulator mRNA translation [22]. The decreased expression of tumor suppressor gene PTEN, a direct antagonist, has been shown to be correlated with poor outcomes in CRC [23].

In Indonesia, there has been limited investigation into the genetic mutation profiles of CRC patients, particularly those with LS. As reported in our previous study and others, several clinical features are associated with this syndrome, including tumor location of which 60-70% found in the right-sided (proximal) colon [6, 10]. In this study we examine the association of oncogenic mutations of KRAS and PIK3CA with MSI, MLH1 promoter methylation and LS as well as the demography and clinicopathology profile of CRC patients in Yogyakarta, Indonesia.

## MATERIALS AND METHODS

### Ethical Statements

This study was approved by the Medical and Health Research Ethics Committee (MHREC) Faculty of Medicine, Public Health, and Nursing of Universitas Gadjah Mada, Yogyakarta (Ethical Approval Number KE/FK/0837/EC/2022). The informed consents have been obtained for the use of tumor samples, clinical data and any other relevant data in the research for all subjects.

### CRC Clinical Sample

For this observational cross-sectional study, we collected 288 CRC samples from the Department of Anatomic Pathology/Dr. Sardjito General Hospital Yogyakarta, Indonesia between 2016 and 2021. Of these, 244 formalin-fixed paraffin-embedded (FFPE) CRC samples were eligible for mutation detection. The patient data acquired for each case included sociodemographic (age and sex), tumor pathology (location/site, stage analysis, TNM stage, histologic grade, lymphatic status, morphology, tumor-infiltrating lymphocytes/TIL), and various clinical parameters (hemoglobin/Hb, albumin serum, Eastern Cooperative Oncology Group/ECOG scale, and body mass Index/BMI).

### DNA Extraction

Paraffin blocks from colorectal cancer patients were cut into 6 pieces with 5 μm thickness. Only one piece in one slide continued with DNA extraction. Genomic DNA was extracted using the QIAamp DNA FFPE tissue kit (Qiagen, USA) according to the manufacturer’s protocol. DNA samples were quantified using a NanoDrop™ spectrophotometer (Thermo Scientific, Waltham, MA, USA). Samples of sufficient concentration and quality were adjusted to a concentration of 20 ng/µL for PCR applications.

### Detection of MSI, BRAF, MLH1 and Oncogenes Mutation

All CRC biomarkers were detected using an IVD kit called BioColomelt-Dx manufactured by Biofarma Ltd, Indonesia. BioColomelt-Dx is a PCR-HRM molecular diagnostics kit for screening MSI, MLH1 promoter methylation, and important oncogene mutations of KRAS (exon 2 and 3), BRAF (V600E), PIK3CA (exon 9 and 20) in FFPE DNA samples. MSI, BRAF, MLH1 methylation promoter detections were previously described as N-LyST panel [24]. The N_LyST panel is a detection method for five mononucleotide microsatellite repeats, BRAF^V600E^ mutations, and MLH1 region C promoter methylation status. For MSI analysis, samples were regarded as MSI if >2 markers (40%) showed instability; otherwise, they were regarded as MSS tumors. Samples showing MSI, BRAF wildtype, and MLH1 promoter methylation (unmethylated) were classified as “Probable Lynch”. Out of the total samples, 244 were successfully determined for oncogene mutation detection, while 223 were suitable for probable Lynch syndrome determination.

### Statistical Analysis

Correlation between variables was calculated using Fisher’s exact test with a significance level of α = 0.05. All analysis was performed using R version 4.3.2.

## RESULTS

### Association between KRAS and PIK3CA Mutation with MSI Status

The analysis of KRAS PIK3CA with MSI status is shown in Table 1. There was a significant association between mutated KRAS and MSI vs MSS (76.19% vs 58.91%; *P*-value=0.038, *P*-value<0.05); and mutated PIK3CA with MSI vs MSS (59.52% vs 40.59%; *P*-value=0.027, *P*-value<0.05). There were N=14/244 samples that had the concomitant mutation of KRAS and BRAF. There was no significant association between KRAS and BRAF concomitant mutation with MSI status (7.14% vs 5.45%; *P*-value= 0.7)

**Table 1.**
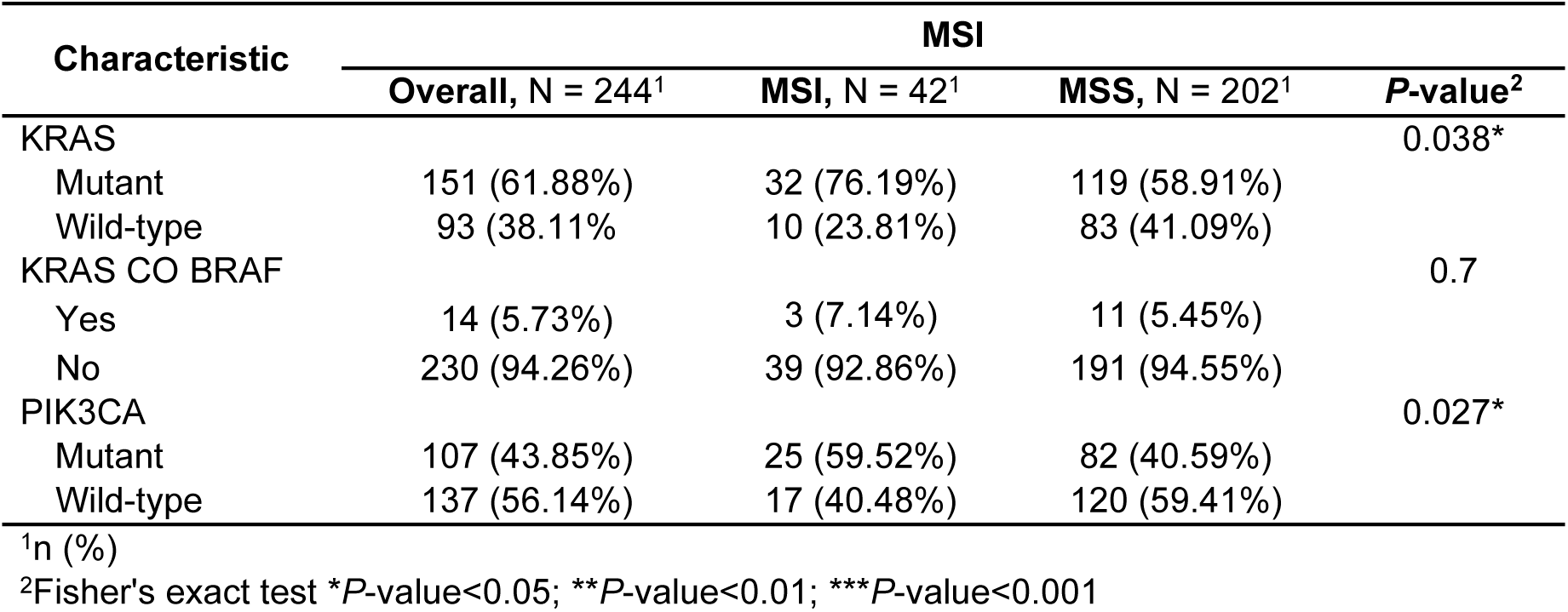
KRAS and PIK3CA with MSI Mutational Status.

### Association between KRAS and PIK3CA Mutation with Probable Lynch Status

The association of KRAS, PIK3CA and Lynch Status showed in Table 2. There was a significant association between mutated KRAS and probable Lynch status (81.25% vs 58.64%; *P*-value=0.018, *P*-value<0.05). However, there were no significant association between PIK3CA and Probable Lynch Status.

**Table 2.**
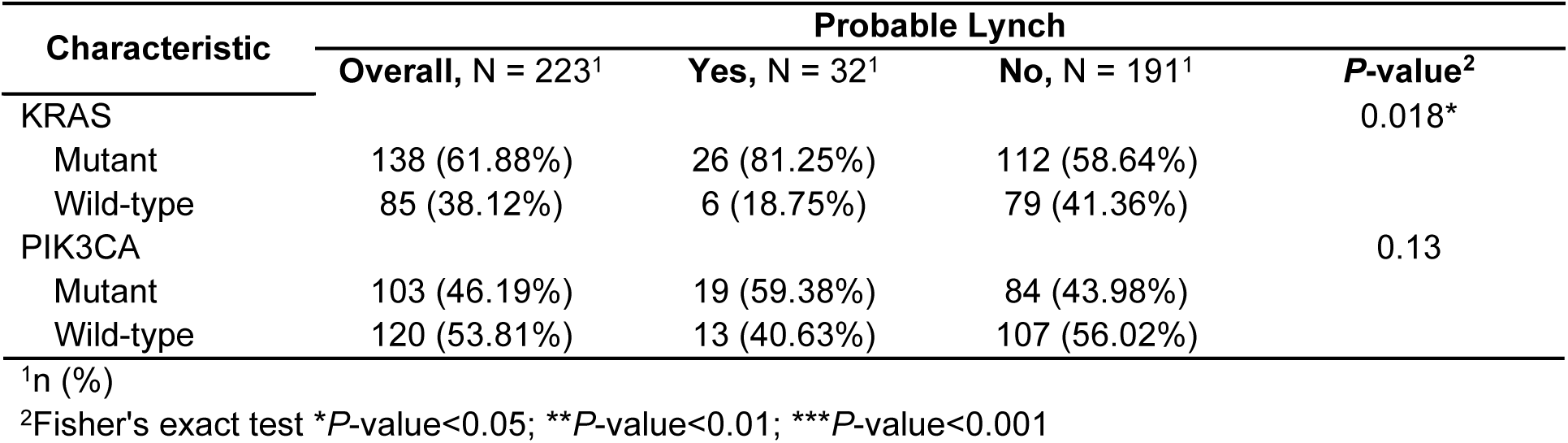
KRAS, PIK3CA and Probable Lynch Status.

### Clinicopathology Association with Oncogene Status

As shown in Table 3, PIK3CA gene mutation frequency was higher in female patients compared to the male patients (57.94% vs 42.06%; *P*-value=0.040, *P*-value<0.05). The lower level of TILs were found in mutated PIK3CA (30.23% vs 16.24%; *P*-value=0.021, *P*-value<0.05). The mutation rate of KRAS in the right sided was higher and statistically significant than the left sided (25.83% vs 13.04%; *P*-value=0.022, *P*-value<0.05). Mutant KRAS was significantly higher in Histology Grade 3 compared to wild-type KRAS (16.44% vs 7.53%; *P*-value=0.038, *P*-value<0.05). There was no significant association between other clinicopathology parameters with KRAS oncogene status.

**Table 3.**
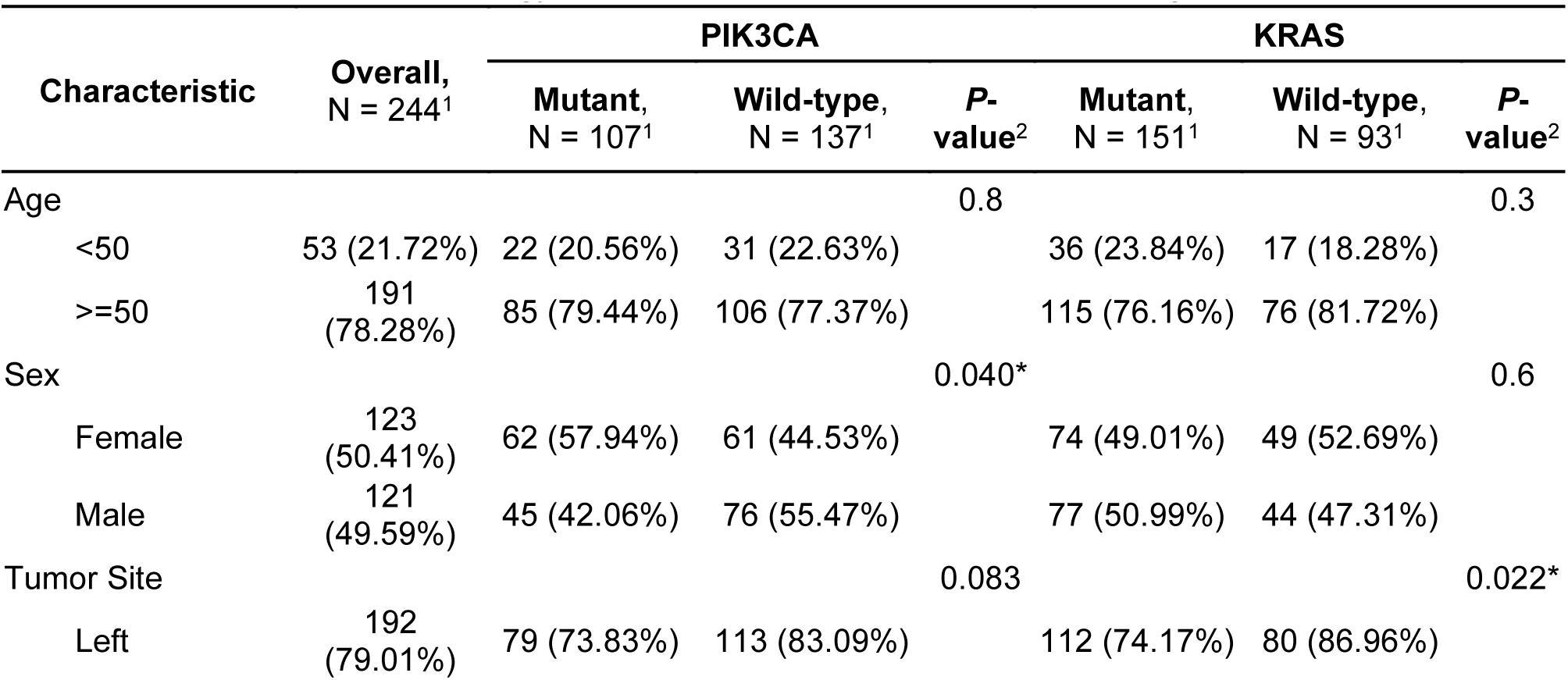
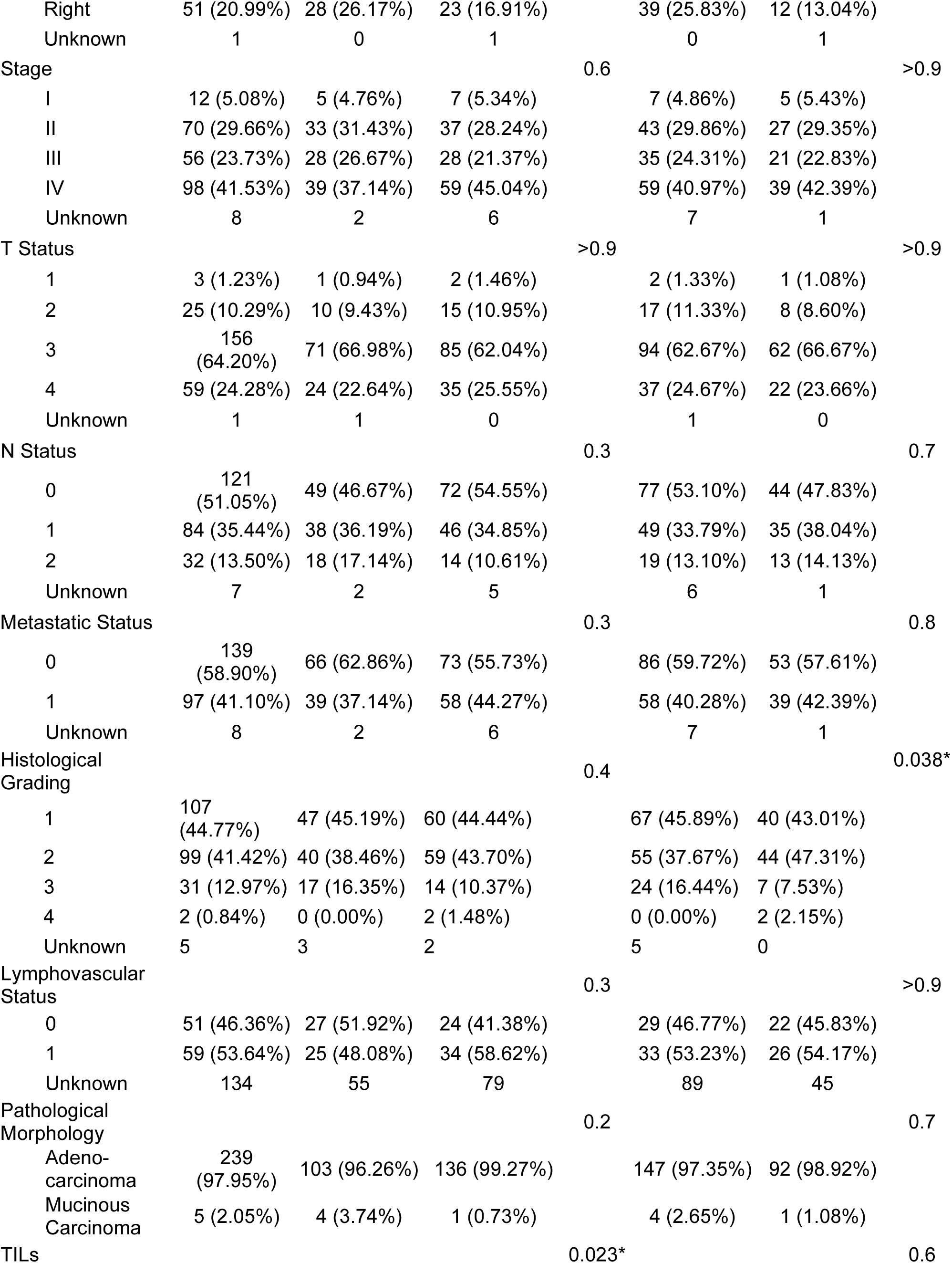
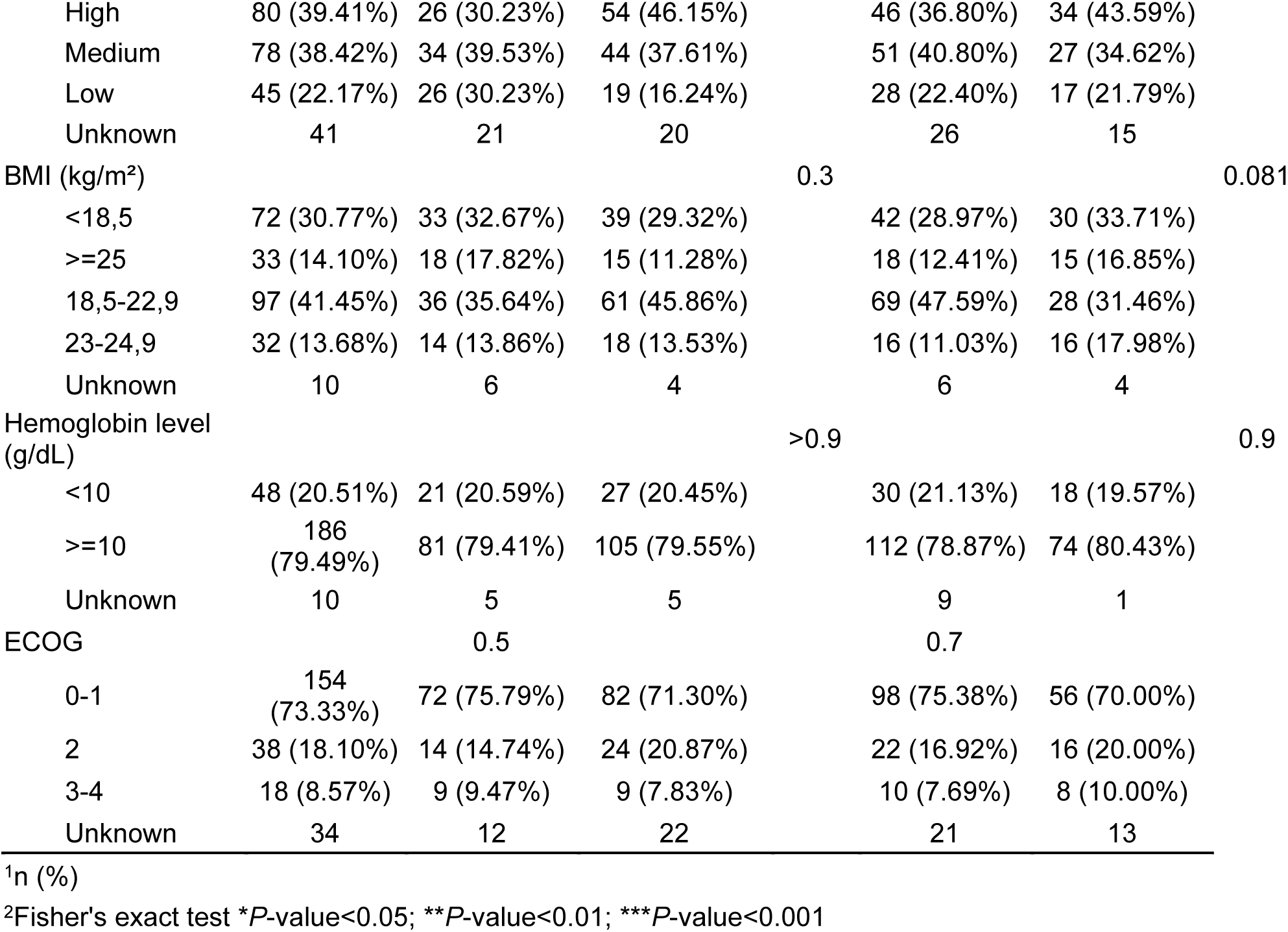
Clinicopathology Association with PIK3CA and KRAS Oncogene Status.

### Overall Survival on KRAS and PIK3CA Mutation Stratified by Lynch Syndrome Status

We analyzed the overall survival based on KRAS and PIK3CA mutations stratified by Lynch syndrome status. The number of samples that met the criteria for this analysis on PIK3CA and KRAS mutation are 220 and 222 samples, respectively. There were no statistically significant differences of overall survival (follow-up period of 120 month) based on KRAS and PIK3CA mutation with Probable Lynch and Non-Lynch status patient (Figure 1).

**Figure 1.**
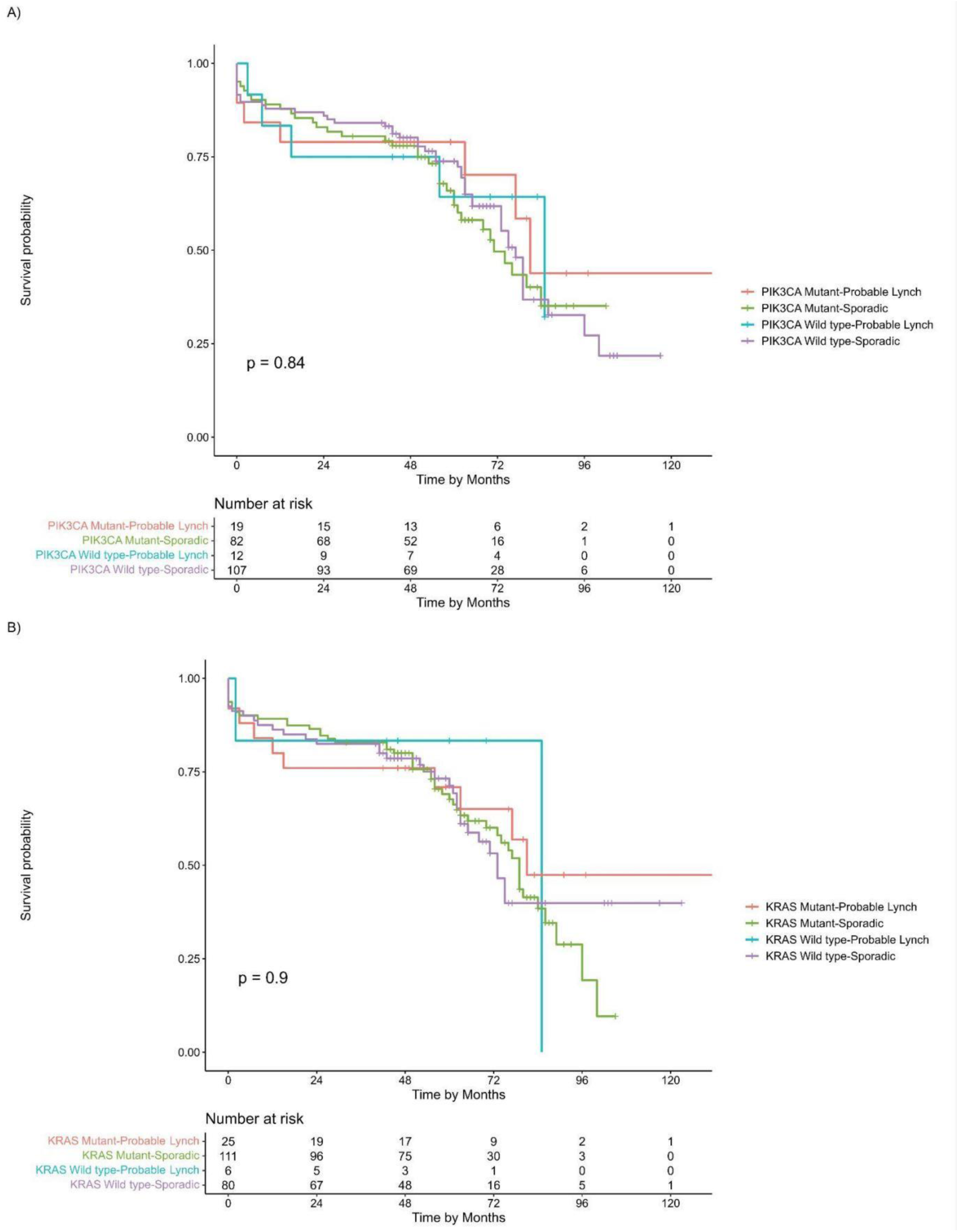
Overall survival curves of CRC patients. A) Survival comparison on PIK3CA mutation and Probable Lynch status; B) Survival comparison on KRAS mutation and Probable Lynch status.

## DISCUSSION

As molecular testing for colorectal cancer (CRC) is not routinely performed in clinical settings, currently there is only limited data on molecular landscape of CRC among the Indonesian patients. We previously reported higher incidence of early onset Colorectal Cancer (EOCRC) and probable Lynch Syndrome (LS) in our patient cohort from Yogyakarta, Indonesia [6]. In this study, we sought to examine the association of oncogenic mutations of KRAS and PIK3CA, LS status and the clinicopathological features.

The frequency of KRAS mutations (exon 2 and exon 3) in our cohort reached 61.88%. This was much higher than what was reported in the Asia-Pacific (37-52%) and Western populations (32-49%) [25–38]. Nevertheless, our data is in line with a previous study in Indonesia, showing that KRAS mutation was found in 71.8% of serrated adenocarcinoma (SA), which is intriguingly higher than the generally reported incidence of 40% [39–41]. Another study, based on NGS (Next Generation Sequencing) analysis, showed that KRAS mutation occurred in 63.6% of 22 Indonesian patients with mostly advanced CRC [42]. Distinct mutations located in codons 13, 14, 34, 58, 59, and 146 were found as opposed to more commonly reported mutations in codons 12, 13, 61, 146 [42–44]. A recent study in China has also reported that KRAS mutation was found in 69.4% of the early lesions [45, 46].

Furthermore, in this study, we found that the KRAS mutation was enriched in MSI compared to MSS cases (76.19% and 58.91%, respectively). We previously reported a lower frequency of BRAF mutations (20.45%) in this MSI-CRC cohort [6], similar to what has been reported in China [47–49]. This is unlike a common dogma in which KRAS mutation is more associated with MSS while BRAF mutation is associated with MSI [50, 51]. We also observed the concomitant KRAS and BRAF mutation on 5.73% of total CRC cases. Despite previously thought as a rare event, there are growing number of studies reporting the co-mutation, this includes a study by Gong et al. (2017) showing the incidence rate of 1.4% of 138 metastatic CRC [52]. Our cohort was highly enriched for metastatic disease (41.1%) which may contribute to the higher frequency of co-mutation. To date, there is no optimal treatment for metastatic CRC harboring both KRAS and BRAF [53]. Consequently, our high co-mutation rate in the Indonesian population can shed new light that warrants further investigation. This study and others have shown that KRAS mutation was more frequent in right sided CRC [37, 54, 55]. The predilection of KRAS mutations for the right side of the colon may be influenced by the fact that the right and left side of the colon have different biology and histopathology in their respective embryological origins [37, 56, 57]. Right-sided CRC often has flat histopathology and a DNA mismatch repair pathway deficiency [58]. This may explain the significant association of mutated KRAS and MSI in our cohort.

Our study reports the PIK3CA (exon 9 and 20) mutation frequency of 43.85%, which is lower compared to a previous study in other regions of Indonesia with smaller sample size, reporting 70.9% mutation [59]. An NGS based study on Indonesian patients with advanced CRC revealed that all patients (100%, n=22) harboring PIK3CA mutation, distinctively in exon 2, 5, 7, 8, 10, 19, and 21, in addition to commonly reported exon 9 and 20 [42]. Similar to our result, a meta-analysis of 44 studies enrolling 17,621 patients has reported that mutations of PIK3CA exon 9 and 20 were associated with MSI, KRAS mutation and right-sided colon [60]. Tumor with MSI is usually associated with a high number of TILs due to the generation of neoantigens hence it serves as a good candidate for immunotherapy [61–63]. However, in this study, we found a subset of lower numbers of TILs in PIK3CA-mutated cases, suggesting an immune-invading mechanism at play.

Interestingly, PD-L1 expression was correlated with PIK3CA mutations, suggesting that cancers with PIK3CA mutations and PD-L1 expression are immunotherapy candidates [64]. Inhibition of the PI3K-AKT pathway may improve effector T cell infiltration in PI3K-altered CRC. Combining PI3K inhibitors with anti-PD-1 could enhance treatment efficacy and CD8+ T cell proliferation [65]. It is also worth noting that PIK3CA mutations are prevalent in the early stages of MSI while it tends to occur later in MSS [66]. These suggest that the combination of PIK3CA mutation, MSI status, and PD-L1 expression could potentially be used to guide treatment selection or prognosis improvement for certain subsets of CRC patients.

As previously reported, our cohort was significantly enriched with EOCRC, defined as <50 years old [6]. We harnessed a robust, simple and affordable test called N_LyST to screen for probable LS, which fit into resource limited settings in Indonesia [24]. The test is a PCR (Polymerase Chain Reaction)-HRMA (High Resolution Melting Analysis) based method, consisting of 5 mononucleotide markers for MSI, MLH1 promoter methylation and BRAF V600E mutation in a single PCR run. We found that there was a high proportion of probable LS in our Indonesian CRC cohort (13.85%) and it was strongly associated with EOCRC, despite there still substantial numbers of EOCRC that were considered as non-LS/ sporadic cases [6].

Similar to other studies, this study found no significant association between oncogenic KRAS and PIK3CA mutations and EOCRC [67]. Nonetheless, in this study, we found high KRAS and PIK3CA mutations, 81.25% and 59.38% respectively, in patients with probable-LS status. These are much higher than previously reported in the literature, showing that KRAS mutations in the LS population ranging between 27-40% [68–72]. We have reported the positive association of PIK3CA mutation with KRAS mutation [73]. There was a statistically significant association between probable LS status and KRAS mutation, but not PIK3CA mutation. It has been reported in the literature that KRAS mutation was more frequently found in LS-related MSI CRC as compared to sporadic MSI CRC [69, 72, 74, 75]. On the other hand, PIK3CA mutation was reported to be more common in somatically mutated MMR-deficient CRC [76].

We observed no differences in overall patients’ survival over a follow-up period of 120-months, between mutant and wildtype subgroups based on probable LS and non-LS/sporadic status for both KRAS and PIK3CA. Prior studies have shown that LS patients with CRC have a better prognosis than those with sporadic CRC, arguably due to its association with MSI and neoantigen generation in enhancing the immune response [77]. Although, as reported previously, we did not see this benefit of survival in our cohort [6]. It is appealing to speculate that high occurrences of KRAS mutation in our probable-LS patients outweigh the survival benefit of MSI. To our knowledge, there have been no studies yet that examine the association of survival rates in KRAS and PIK3CA mutations with LS in CRC.

Lynch syndrome is highly heterogeneous, showing high variability in age at onset (despite enriched in EOCRC), penetrance of cancer, and clinical presentations, which may be partly attributable to the molecular profiles of carcinomas. As reviewed by Helderman, et al. (2021), LS heterogeneity is attributable to a variety of different molecular pathways of tumor development, and only partly depends on which MMR gene is mutated [78]. It is now recognized that LS CRCs develop via one of three pathways. The first pathway is adenoma-carcinoma pathway, in which adenomas develop independently of MMR deficiency. The second and third pathway is MMR-deficient crypt foci (MMR-DCF)–Adenoma–Carcinoma pathway and MMR-DCF–Carcinoma pathway, which both start with MMR deficiency and is either followed by adenoma formation or results directly in a carcinoma [78, 79]. As widely known, APC mutations are more closely related to the development of adenomas, while CTNNB1 mutations appear to be associated with the MMR-DCF–Carcinoma pathway [74, 78].

Understanding the molecular landscape of LS, such as RAF/MEK/ERK and PI3K/PTEN/AKT signaling, will allow more detailed stratification of LS patients and will facilitate the provision of optimal care to each patient, including the diagnosis, surveillance, and treatment. For instance, activating PIK3CA variants are potentially susceptible to preventive aspirin therapy by inducing the transcription of COX2 gene increasing the production of PGE2 [74, 80]. This is in addition to known resistance of anti-EGFR of CRC harboring KRAS, BRAF and PIK3CA mutations [81, 82]. However, it is not yet known whether cancers that develop during aspirin therapy have a specific molecular signature [83].

In summary, despite the limitations of this study, including being conducted in a single center and utilizing a low throughput (PCR-based) workflow for mutation detections, this study has contributed to the better understanding of CRC molecular features in an underrepresented population in current global literatures. Our initial findings described in this paper and previous reports from our and other studies have indicated distinct genetic make-up such as high probability of LS, high frequency KRAS, and PIK3CA mutations, and lower BRAF mutation among CRC in Indonesia. This may underpin its unique clinical characteristics such as higher number of young patients and advanced disease stage. Further comprehensive multicenter analysis using high throughput techniques such as NGS is important to provide a more complete picture of the CRC carcinogenesis in Indonesia, with particular emphasis on EOCRC and LS. Understanding the global molecular landscape of CRC may reveal new knowledge that could challenge the current dogma, hence improving the effort to provide better care for the disease.

## CONCLUSION

The high probability of LS in Indonesian CRC patients is associated with KRAS and PIK3CA mutations. It improved our understanding of CRC molecular features in an underrepresented population in global literature.

## Acknowledgments

The authors thank Aru W Sudoyo and Ahmad Utomo for critical input into study design and analysis of results, Yasjudan Rastrama Putra and Yana Suryani for technical assistance and coordination.

## Informed Consent

All subjects provided written informed consent.

## Author Contributions

Conception or design of the work: D.S.H., S.H.H., and S.S.;

Methodology: D.S.H., N.Y., G.A., A.M.R., and S.H.H.;

Software: G.A., H.H., M.Z.A., A.M.R., W.K.; and S.S.;

Validation: D.S.H., N.Y., G.A., A.M.R., S.H.H., and S.S.;

Formal Analysis: D.S.H., N.Y., G.A., H.H., M.Z.A., A.M.R., S.H.H., and S.S.;

Investigation: N.Y., G.A., H.H., A.M.R. W.K.; and S.S.;

Resources: D.S.H., N.Y., A.Y.H., J.K., S.H.H., and S.S.;

Data Curation: D.S.H., N.Y., G.A., V.L., H.H., Z.H., A.N.G., A.M.R, and S.S.;

Writing–Original Draft Preparation: D.S.H., V.L., H.H., Z.H., A.N.G., and S.S.;

Writing–Review and Editing: D.S.H., A.Y.H., M.I., J.K., S.H.H., and S.S.;

Visualization: G.A., M.Z.A., and S.S.;

Supervision: D.S.H., N.Y., M.I., S.H.H. and S.S.;

Project Administration: G.A., V.L., H.H., Z.H., and A.M.R.;

Funding Acquisition: D.S.H., M.I., S.H.H. and S.S.;

Approval of the version to be published: all authors. All authors have read and agreed to the published version of the manuscript.

## Data Availability Statement

All data and related metadata underlying the findings reported in this manuscript have been provided as part of the submitted article. Any additional data that might support the findings of this study are available from the corresponding author, S.S., upon reasonable request.

