## Supplementary material for "High Probability of Lynch Syndrome among colorectal cancer patients in Indonesia is associated with higher occurrence of KRAS and PIK3CA mutations": Table: TABLE.docx

Table 1. KRAS and PIK3CA with MSI Mutational Status

| **Characteristic** | **MSI** | | | |
| --- | --- | --- | --- | --- |
|  | **Overall,** N = 244^a^ | **MSI,** N = 42^a^ | **MSS,** N = 202^a^ | ***P*-value** |
| KRAS |  |  |  | 0.038^b^ |
| Mutant | 151 (61.88%) | 32 (76.19%) | 119 (58.91%) |  |
| Wild-type | 93 (38.11% | 10 (23.81%) | 83 (41.09%) |  |
| KRAS CO BRAF |  |  |  | 0.7 |
| Yes | 14 (5.73%) | 3 (7.14%) | 11 (5.45%) |  |
| No | 230 (94.26%) | 39 (92.86%) | 191 (94.55%) |  |
| PIK3CA |  |  |  | 0.027^b^ |
| Mutant | 107 (43.85%) | 25 (59.52%) | 82 (40.59%) |  |
| Wild-type | 137 (56.14%) | 17 (40.48%) | 120 (59.41%) |  |

Values are presented as number (%).

^a^Number of samples.

^b^Fisher's exact test *P*-value<0.05.

MSI, Microsatellite Instability; MSS, Microsatellite Stability.

### Table 2. KRAS, PIK3CA and Probable Lynch Status

| **Characteristic** | **Probable Lynch** | | | |
| --- | --- | --- | --- | --- |
|  | **Overall,** N = 223^a^ | **Yes,** N = 32^a^ | **No,** N = 191^a^ | ***P*-value** |
| KRAS |  |  |  | 0.018^b^ |
| Mutant | 138 (61.88%) | 26 (81.25%) | 112 (58.64%) |  |
| Wild-type | 85 (38.12%) | 6 (18.75%) | 79 (41.36%) |  |
| PIK3CA |  |  |  | 0.13 |
| Mutant | 103 (46.19%) | 19 (59.38%) | 84 (43.98%) |  |
| Wild-type | 120 (53.81%) | 13 (40.63%) | 107 (56.02%) |  |

Values are presented as number (%).

^a^Number of samples.

^b^Fisher's exact test *P*-value<0.05.

Table 3. Clinicopathology Association with PIK3CA and KRAS Oncogene Status

| **Characteristic** | **Overall,**  N = 244^a^ | **PIK3CA** | | | **KRAS** | | |
| --- | --- | --- | --- | --- | --- | --- | --- |
|  |  | **Mutant**,  N = 107^a^ | **Wild-type**,  N = 137^a^ | ***P*-value** | **Mutant**,  N = 151^a^ | **Wild-type**,  N = 93^a^ | ***P*-value** |
| Age |  |  |  | 0.8 |  |  | 0.3 |
| <50 | 53 (21.72%) | 22 (20.56%) | 31 (22.63%) |  | 36 (23.84%) | 17 (18.28%) |  |
| ≥ 50 | 191 (78.28%) | 85 (79.44%) | 106 (77.37%) |  | 115 (76.16%) | 76 (81.72%) |  |
| Sex |  |  |  | 0.040^b^ |  |  | 0.6 |
| Female | 123 (50.41%) | 62 (57.94%) | 61 (44.53%) |  | 74 (49.01%) | 49 (52.69%) |  |
| Male | 121 (49.59%) | 45 (42.06%) | 76 (55.47%) |  | 77 (50.99%) | 44 (47.31%) |  |
| Tumor Site |  |  |  | 0.083 |  |  | 0.022^b^ |
| Left | 192 (79.01%) | 79 (73.83%) | 113 (83.09%) |  | 112 (74.17%) | 80 (86.96%) |  |
| Right | 51 (20.99%) | 28 (26.17%) | 23 (16.91%) |  | 39 (25.83%) | 12 (13.04%) |  |
| Unknown | 1 | 0 | 1 |  | 0 | 1 |  |
| Stage |  |  |  | 0.6 |  |  | >0.9 |
| I | 12 (5.08%) | 5 (4.76%) | 7 (5.34%) |  | 7 (4.86%) | 5 (5.43%) |  |
| II | 70 (29.66%) | 33 (31.43%) | 37 (28.24%) |  | 43 (29.86%) | 27 (29.35%) |  |
| III | 56 (23.73%) | 28 (26.67%) | 28 (21.37%) |  | 35 (24.31%) | 21 (22.83%) |  |
| IV | 98 (41.53%) | 39 (37.14%) | 59 (45.04%) |  | 59 (40.97%) | 39 (42.39%) |  |
| Unknown | 8 | 2 | 6 |  | 7 | 1 |  |
| Tumor Status |  |  |  | >0.9 |  |  | >0.9 |
| 1 | 3 (1.23%) | 1 (0.94%) | 2 (1.46%) |  | 2 (1.33%) | 1 (1.08%) |  |
| 2 | 25 (10.29%) | 10 (9.43%) | 15 (10.95%) |  | 17 (11.33%) | 8 (8.60%) |  |
| 3 | 156 (64.20%) | 71 (66.98%) | 85 (62.04%) |  | 94 (62.67%) | 62 (66.67%) |  |
| 4 | 59 (24.28%) | 24 (22.64%) | 35 (25.55%) |  | 37 (24.67%) | 22 (23.66%) |  |
| Unknown | 1 | 1 | 0 |  | 1 | 0 |  |
| Node Status |  |  |  | 0.3 |  |  | 0.7 |
| 0 | 121 (51.05%) | 49 (46.67%) | 72 (54.55%) |  | 77 (53.10%) | 44 (47.83%) |  |
| 1 | 84 (35.44%) | 38 (36.19%) | 46 (34.85%) |  | 49 (33.79%) | 35 (38.04%) |  |
| 2 | 32 (13.50%) | 18 (17.14%) | 14 (10.61%) |  | 19 (13.10%) | 13 (14.13%) |  |
| Unknown | 7 | 2 | 5 |  | 6 | 1 |  |
| Metastatic Status |  |  |  | 0.3 |  |  | 0.8 |
| 0 | 139 (58.90%) | 66 (62.86%) | 73 (55.73%) |  | 86 (59.72%) | 53 (57.61%) |  |
| 1 | 97 (41.10%) | 39 (37.14%) | 58 (44.27%) |  | 58 (40.28%) | 39 (42.39%) |  |
| Unknown | 8 | 2 | 6 |  | 7 | 1 |  |
| Histological Grading |  |  |  | 0.4 |  |  | 0.038^b^ |
| 1 | 107 (44.77%) | 47 (45.19%) | 60 (44.44%) |  | 67 (45.89%) | 40 (43.01%) |  |
| 2 | 99 (41.42%) | 40 (38.46%) | 59 (43.70%) |  | 55 (37.67%) | 44 (47.31%) |  |
| 3 | 31 (12.97%) | 17 (16.35%) | 14 (10.37%) |  | 24 (16.44%) | 7 (7.53%) |  |
| 4 | 2 (0.84%) | 0 (0.00%) | 2 (1.48%) |  | 0 (0.00%) | 2 (2.15%) |  |
| Unknown | 5 | 3 | 2 |  | 5 | 0 |  |
| Lymphovascular Status |  |  |  | 0.3 |  |  | >0.9 |
| 0 | 51 (46.36%) | 27 (51.92%) | 24 (41.38%) |  | 29 (46.77%) | 22 (45.83%) |  |
| 1 | 59 (53.64%) | 25 (48.08%) | 34 (58.62%) |  | 33 (53.23%) | 26 (54.17%) |  |
| Unknown | 134 | 55 | 79 |  | 89 | 45 |  |
| Pathological Morphology |  |  |  | 0.2 |  |  | 0.7 |
| Adenocarcinoma | 239 (97.95%) | 103 (96.26%) | 136 (99.27%) |  | 147 (97.35%) | 92 (98.92%) |  |
| Mucinous Carcinoma | 5 (2.05%) | 4 (3.74%) | 1 (0.73%) |  | 4 (2.65%) | 1 (1.08%) |  |
| TILs |  |  |  | 0.023^b^ |  |  | 0.6 |
| High | 80 (39.41%) | 26 (30.23%) | 54 (46.15%) |  | 46 (36.80%) | 34 (43.59%) |  |
| Medium | 78 (38.42%) | 34 (39.53%) | 44 (37.61%) |  | 51 (40.80%) | 27 (34.62%) |  |
| Low | 45 (22.17%) | 26 (30.23%) | 19 (16.24%) |  | 28 (22.40%) | 17 (21.79%) |  |
| Unknown | 41 | 21 | 20 |  | 26 | 15 |  |
| BMI (kg/m²) |  |  |  | 0.3 |  |  | 0.081 |
| <18,5 | 72 (30.77%) | 33 (32.67%) | 39 (29.32%) |  | 42 (28.97%) | 30 (33.71%) |  |
| >=25 | 33 (14.10%) | 18 (17.82%) | 15 (11.28%) |  | 18 (12.41%) | 15 (16.85%) |  |
| 18,5-22,9 | 97 (41.45%) | 36 (35.64%) | 61 (45.86%) |  | 69 (47.59%) | 28 (31.46%) |  |
| 23-24,9 | 32 (13.68%) | 14 (13.86%) | 18 (13.53%) |  | 16 (11.03%) | 16 (17.98%) |  |
| Unknown | 10 | 6 | 4 |  | 6 | 4 |  |
| Hemoglobin level (g/dL) |  |  |  | >0.9 |  |  | 0.9 |
| <10 | 48 (20.51%) | 21 (20.59%) | 27 (20.45%) |  | 30 (21.13%) | 18 (19.57%) |  |
| >=10 | 186 (79.49%) | 81 (79.41%) | 105 (79.55%) |  | 112 (78.87%) | 74 (80.43%) |  |
| Unknown | 10 | 5 | 5 |  | 9 | 1 |  |
| ECOG |  | 0.5 |  |  | 0.7 |  |  |
| 0-1 | 154 (73.33%) | 72 (75.79%) | 82 (71.30%) |  | 98 (75.38%) | 56 (70.00%) |  |
| 2 | 38 (18.10%) | 14 (14.74%) | 24 (20.87%) |  | 22 (16.92%) | 16 (20.00%) |  |
| 3-4 | 18 (8.57%) | 9 (9.47%) | 9 (7.83%) |  | 10 (7.69%) | 8 (10.00%) |  |
| Unknown | 34 | 12 | 22 |  | 21 | 13 |  |

Values are presented as number (%)

^a^Number of samples

^b^Fisher's exact test *P*-value<0.05

BMI, Body Mass Index; ECOG, Eastern Cooperative Oncology Group; TILs, Tumor Infiltrating Lymphocytes.
